# Lack of benefit of temozolomide for *MGMT* methylated patients with high vascular glioblastoma: a confirmatory study

**DOI:** 10.1101/2021.09.01.21262673

**Authors:** María del Mar Álvarez-Torres, Elies Fuster-García, Carmen Balaña, Josep Puig, Juan Miguel García-Gómez

**Affiliations:** Universitat Politècnica de València, ITACA, Biomedical Data Science Laboratory. Valencia, Spain; Oslo University Hospital, Department of Diagnostic Physics. Oslo, Norway; Institut Catala Oncologia Badalona. Barcelona, Spain; Institut de Diagnostic per la Image (IDI), Hospital Dr Josep Trueta. Girona, Spain

**Keywords:** Glioblastoma, *MGMT* methylation, tumor vascularity, chemotherapy, adjuvant temozolomide, temozolomide cycles, MRI perfusion, rCBV, survival, personalized medicine

## Abstract

In this study we evaluated the benefit on survival of the combination of *MGMT* methylation and moderate vascularity in glioblastoma using a retrospective dataset of 123 patients from a multicenter cohort. MRI processing and calculation of relative cerebral blood volume (rCBV), used to define moderate- and high-vascular groups, were performed with the automatic ONCOhabitats method. We assessed the previously proposed rCBV threshold (10.7) and the new calculated ones (9.1 and 9.8) to analyze the association with survival for different populations according to vascularity and *MGMT* methylation status. We found that patients included at the moderate-vascular group had longer survival when *MGMT* is methylated (significant median survival difference of 174 days, p = 0.0129^*^). However, we did not find significant differences depending on the *MGMT* methylation status for the high-vascular group (p = 0.9119). In addition, we investigated the combined correlation of *MGMT* methylation status and rCBV with the prognostic effect of the number of temozolomide cycles, and only significant results were found for the moderate-vascular group. In conclusion, there is a lack of benefit of temozolomide for *MGMT* methylated patients with high vascular glioblastomas. Preliminary results suggest that patients with moderate vascularity and methylated *MGMT* would benefit more from prolonged adjuvant chemotherapy.

**Simple Summary:** Despite the complete treatment with surgery, chemotherapy and radiotherapy, patients with glioblastoma have a devasting prognosis. Although the role of extending temozolomide treatment has been explored, the results are inconclusive. Recent evidence suggested that tumor vascularity may be a modulating factor in combination with *MGMT* methylation on the effect of temozolomide-based therapies, opening new possibilities for personalized treatments. Before proposing a prospective interventional clinical study, it is necessary to confirm the beneficial effect of the combined effect of *MGMT* methylation and moderate tumor vascularity. As well as the lack of benefit of temozolomide in patients with a highly vascular tumor.

## 1. Introduction

Glioblastoma patients remain a devastating prognosis of 12-15 months from diagnosis [1, 2] despite an intrusive treatment including tumor resection, radiotherapy, and concomitant and maintenance chemotherapy with temozolomide [3]. This standard treatment, proposed by Stupp in 2005 [3] was demonstrated to be the most effective in terms of overall survival but, due to strong interpatient heterogeneity, it is not equally efficient for all patients [4].

Several studies have evaluated the efficacy of this treatment depending on several conditions as extend of tumor resection [5-11], age [12], the methylation of the O6-methylguanine-DNA methyltransferase (MGMT) promoter gene [13], dose of temozolomide [14-20], the addition of new agents [21-23] or the device tumor treating fields [24-30], that in fact is the only modification that has proven to increase survival.

The optimal number of cycles of temozolomide in the maintenance phase has also been a matter of debate [31, 32]. This is due to the heterogeneity of uses or interpretation of the term ‘maintenance’ or ‘adjuvant therapy’ in a disease as glioblastoma was surgery seldom achieves a complete resection. The number of cycles administered is clearly variable in the clinical setting or even in the different trials [33]. The only prospective trial assessing the role of extending temozolomide further than six cycles is a randomized phase II trial that did not demonstrate differences in progression-free survival or overall survival [14]. The European Association of Neuro-oncology guidelines recommend six cycles of maintenance therapy [34].

A same treatment for all patients with glioblastoma has been demonstrated ineffective. The availability of robust markers to characterize interpatient heterogeneity, and therefore, to discriminate different subgroups could lead to a more personalized medicine approach. In this line, imaging markers derived from MRI and combined with the capabilities of artificial intelligence can provide individually specific variations of the Stupp treatment. This would allow better prognosis and facilitate the clinical decision-making for patient treatment in a non-invasive way and not-extra cost [35-39].

Currently, glioma classification, decision making, and management of glioblastoma are still based on molecular biomarkers [1, 40-43]. One of the most relevant biomarkers, related with the Stupp treatment efficacy, is MGMT status [44], present in approximately 50% of glioblastomas [45]. This alteration affects the ability to repair DNA damaged induced by alkylating agents such as temozolomide, allowing for a more durable and efficient effect of chemotherapy [44, 46].

A recent study with a multicenter cohort of 96 glioblastoma patients [47] concluded that MGMT methylation may benefit overall survival only in patients with moderately vascularized glioblastomas, defined by MRI perfusion–based marker, such as relative cerebral blood volume (rCBV). This study opened the possibility of investigating vascularity as a determinant factor, in combination with methylation status, on the benefit of temozolomide cycles.

In this study, we aimed to evaluate the combined effect of MGMT methylation and tumor vascularity on patient survival assessing the performance of the proposed rCBV threshold for patient stratification. We also assessed the implications of the association between MGMT methylation and vascularity on the benefit of the number of administered temozolomide cycles in different group of glioblastoma patients.

## 2. Materials and Methods

### 2.1. Patient Information

For this study, 123 glioblastoma patients were included from the GLIOCAT database [48], which includes patients from the following 6 centers from Cataluña, Spain: (I) Instituto Catalán de Oncología (ICO) de Badalona (Barcelona), (II) Hospital del Mar (Barcelona), (III) Hospital Clínic (Barcelona), (IV) ICO Hospitalet (Barcelona), (V) ICO Girona (Girona) and, (VI) Hospital Sant Pau (Barcelona).

A Material Transfer Agreement was approved by all the participating centers and ethical approval was given by: the Ethical Committee of Instituto Catalán de Oncología de Badalona, the Ethical Committee of Hospital del Mar in Barcelona, the Ethical Committee of Hospital Clínic of Barcelona, the Ethical Committee of ICO Hospitalet in Barcelona, the Ethical Committee of ICO Girona and, the Ethical Committee of Hospital Sant Pau in Barcelona.

The inclusion criteria were: (a) adult patients (age >18 years) with histopathological confirmation of glioblastoma; diagnosed between June 2007 – May 2015, (b) with access to the preoperative MRI studies, including: pre- and post-gadolinium T1-weighted, T2-weighted, Fluid-Attenuated Inversion Recovery (FLAIR) and Dynamic Susceptibility Contrast (DSC) T2^*^-weighted perfusion sequences; (c) with *MGMT* methylation status information, (d) with a minimum survival of 30 days and, (e) with tumor resection.

Patients still alive at readout were considered censored observations. The date of censorship was the last date of contact with the patient or, if not available, the date of the last MRI exam.

### 2.2. Magnetic Resonance Imaging

Standard-of-care MR examinations were obtained for each patient before surgery, including pre- and post-gadolinium-based contrast agent enhanced T1-weighted MRI, as well as T2-weighted, FLAIR T2-weighted, and DSC T2^*^ perfusion MRI.

### 2.3. MRI Processing and Vascular Marker Calculation

To process the MRIs and calculate the imaging vascular markers, we used the Hemodynamic Tissue Signature (HTS) method [49, 50], freely accessible at the ONCOhabitats platform at www.oncohabitats.upv.es. The HTS is an automated unsupervised method developed to describe the heterogeneity of the enhancing tumor and edema tissues at morphological and vascular levels, and to calculate robust biomarkers with prognostic and patient stratification capabilities. This method includes the following four phases (Figure 1):

**Figure 1:**
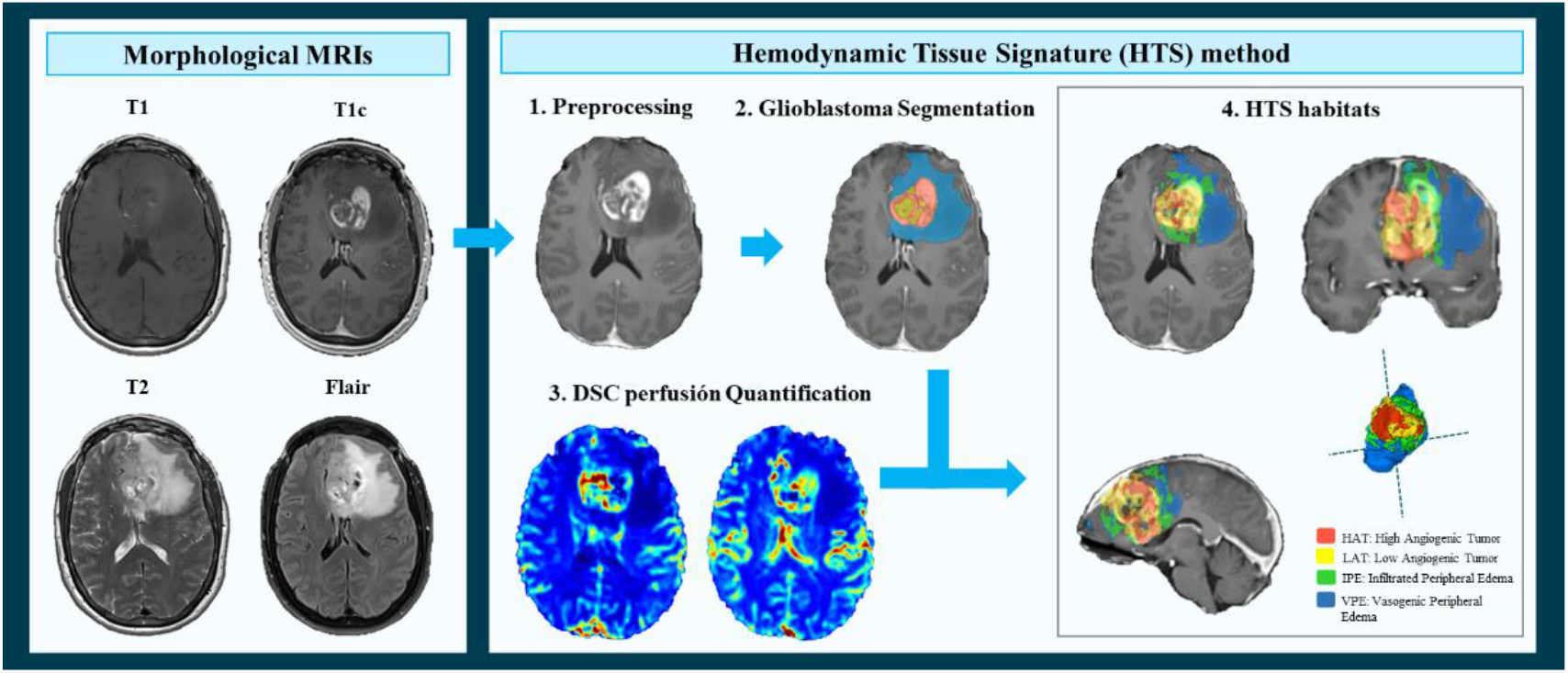
Hemodynamic Tissue Signature (HTS) method, including the four phases: 1. Preprocessing of morphological MRIs (T1, T1c, T2 and Flair); 2. Glioblastoma tissue segmentation; 3.DSC perfusion quantification and, 4. HTS vascular habitats: High Angiogenic Tumor (HAT), Low Angiogenic Tumor (LAT), Infiltrated Peripheral Edema (IPE) and Vasogenic Peripheral Edema (VPE).

1. MRI Pre-processing. This phase includes voxel isotropic resampling of all MR images, correction of the magnetic field in homogeneities and noise, rigid intra-patient MRI registration, and skull-stripping.
2. Glioblastoma tissue segmentation. It is performed by using an unsupervised segmentation method, which implements a state-of-the-art deep-learning 3D convolutional neural network (CNN), which takes as input the T1c, T2, and Flair MRIs. This method is based on Directional Class Adaptive Spatially Varying Finite Mixture Model, or DCA-SVFMM, which consists of a clustering algorithm that combines Gaussian mixture modeling with continuous Markov random fields to take advantage of the self-similarity and local redundancy of the images.
3. DSC perfusion quantification. In this phase, biomarkers such as the relative cerebral blood volume (rCBV) maps, as well as relative cerebral blood flow (rCBF) or Mean Transit Time (MTT), are calculated for each patient. T1-weighted leakage effects are automatically corrected using the Boxerman method [51], while gamma-variate curve fitting is employed to correct for T2 extravasation phase. rCBV maps are calculated by numerical integration of the area under the gamma-variate curve. The Arterial Input Function (AIF) is automatically quantified with a divide and conquer algorithm.
4. Hemodynamic Tissue Signature and Vascular Habitats. The HTS provides an automated unsupervised method to describe the heterogeneity of the enhancing tumor and edema tissues, in terms of the angiogenic process located at these regions. We consider 4 sub-compartments for the glioblastoma, two within the active tumor: High Angiogenic Tumor habitat (HAT) and Low Angiogenic Tumor habitat (LAT); and two within the edema: Infiltrated Peripheral Edema habitat (IPE) and Vasogenic Peripheral Edema habitat (VPE). These four habitats are obtained by means of a DCA-SVFMM structure clustering of rCBV and rCBF maps. The clustering consists of two stages: (a) a two-class clustering of the whole enhancing tumor and edema ROIs and (b) a two-class clustering performed by using only the rCBV and rCBF data within the ROIs obtained in stage a.

A more detailed description of the methodology is included in [49, 50]. In addition, the HTS method and the vascular biomarkers were validated in an international multicenter study and results were published in [52].

To validate the combined effect of MGMT methylation and vascularity, we used the maximum relative cerebral blood volume (rCBVmax) calculated in the HAT habitat, since it is shown to be the most relevant prognostic marker calculated with the HTS method [50, 52, 53] and it was used in the previous study to define the vascular groups [47].

### 2.4. Moderate- and High-vascular Groups

The entire cohort was divided in two groups according to the tumor vascularity: the moderate-vascular group and the high-vascular group. To determine these groups, we carried out the analysis independently using three different thresholds (th) of the HAT rCBVmax:

I. The threshold proposed in the literature (49) (th = 10.7). It was calculated as the median rCBVmax of 96 patients included in an international multicenter study.
II. The median rCBVmax of the current study cohort (th = 9.1). Calculated from the 123 patients included in the present study.
III. The combined threshold of both cohorts (th = 9.8). It is calculated considering the 219 patients from two independent multicenter studies.

The purpose to evaluate these three different thresholds is to validate the previous results and the threshold proposed in the literature [47] with an independent multicenter cohort; but also to analyze the stratification capability of the HAT rCBVmax when using the specific threshold calculated from the current study cohort. Finally, proposing a combined threshold calculated from both independent cohorts with 219 patients will allow most reproducible results.

### 2.5. Statistical Analyses

#### 2.5.1. Dataset description: differences between methylated and unmethylated *MGMT* groups

We described the main demographic, clinical, and molecular variables for the entire cohort and for methylated and unmethylated *MGMT* populations. The analyzed variables for each population were: gender, age at diagnosis, survival times, extent of tumor resection, completeness of concomitant chemotherapy, number of adjuvant temozolomide cycles, *IDH1* mutation status and, rCBV_max_ at HAT habitat. Possible differences in the distributions of these variables for the populations with methylated and unmethylated *MGMT* were assessed using Mann-Whitney U test (for ordinal or continuous variables) or Fisher exact test (for nominal variables) in MATLAB R2017b (MathWorks, Natick, MA). The significance level used in all the statistical analyses was 0.05.

#### 2.5.2. Association between *MGMT* methylation, tumor vascularity and patient survival

To validate the previous results published in [47], which showed a significant correlation between *MGMT* methylation status with overall survival only for those patients with moderate vascularized tumors, we carried out the Uniparametric Cox proportional hazard regression. These analyses were carried out for the entire cohort, and independently for the methylated and unmethylated *MGMT* groups and using the three studied thresholds. The proportional hazard ratios (HRs) with a 95% confidence interval (CI), as well as the associated p-values are reported.

Kaplan Meier test was carried out to evaluate the different effect on survival of *MGMT* methylation status, depending on tumor vascularity and, the Log rank was used to determine any statistical differences between the estimated survival functions of the different *MGMT* methylation populations, both at moderate- and high-vascular groups. The number of patients included in each group, the median OS rates of each group, the differential OS and the p-values are reported.

The following results were carried out using the (III) combined threshold (th = 9.8), since authors consider it as the most robust threshold because its calculation was derived from data of 214 patients from two different multicenter datasets and could generate more repeatable results.

#### 2.5.3. Benefit of adjuvant temozolomide cycles in different groups of glioblastoma patients

To analyze the combined effect of MGMT methylation and the number of adjuvant temozolomide cycles on survival, a Multiparametric Cox regression analysis were carried out including *MGMT* methylation status and number of temozolomide cycles for the entire cohort and, independently for the moderate- and high vascular groups. The number of temozolomide cycles was a continuous variable with a minimum of 0 to a maximum of 12 cycles.

In addition, to study differences in patient survival associated with the number of administered temozolomide cycles, a boxplot was carried out for each group (defined by *MGMT* methylation status and tumor vascularity).

## 3. Results

### 3.1. Study cohort

This study includes data from 123 patients with primary glioblastoma. Table 1 summarizes the main demographic, clinical and biological characteristics of the entire cohort and independently for the groups of patients with methylated and unmethylated *MGMT*. In addition, p-values derived from Mann-Whitney U test or Fisher exact test are included.

**Table 1:** Demographic, clinical and biological characteristics of the entire cohort, and the groups with methylated and unmethylated MGMT. P-values derived from Mann-Whitney (MW) test or Fisher exact (FE) test analyzing differences between methylated and unmethylated *MGMT*.

| Variables | Entire cohort | Methylated<br>MGMT<br>population | Unmethylated<br>MGMT<br>population | P-values<br>(MW/FE) |
| --- | --- | --- | --- | --- |
| **Number of patients** | 123 | 67 | 56 | - |
| **Gender** |  |  |  |  |
| - % females | 41.5 | 43.3 | 39.2 | 0.7150 |
| **Age at diagnosis (years)** |  |  |  | 0.8973 |
| - Mean | 60 | 62 | 58 |  |
| - Range | [32,80] | [33,80] | [32,77] |  |
| **Overall Survival (months)** |  |  |  | 0.1214 |
| - Mean | 20.2 | 22.4 | 17.6 |  |
| - Median | 17.1 | 19.3 | 15.5 |  |
| - Range | [2.7,72.8] | [2.7,71.6] | [2.7,72.8] |  |
| **Extent of Resection (#patients)** |  |  |  | 0.4524 |
| - Complete | 45 | 27 | 18 |  |
| - Partial | 78 | 40 | 38 |  |
| **Concomitant chemotherapy (#patients)** |  |  |  | 0.3788 |
| -Complete | 110 | 58 | 52 |  |
| -Incomplete | 13 | 9 | 4 |  |
| **Adjuvant chemotherapy (number of cycles)** |  |  |  | 0.4435 |
| - Mean | 4 | 5 | 4 |  |
| - Median | 5 | 5 | 4 |  |
| - Range | [0,12] | [0,12] | [0,12] |  |
| **IDH1 mutation status** |  |  |  | 1.0000 |
| -Mutated | 2 | 1 | 1 |  |
| -Wild type | 93 | 51 | 42 |  |
| -Unknown | 28 | 15 | 13 |  |
| **HAT rCBV<sub>max</sub>** |  |  |  | 0.4150 |
| - Mean | 9.77 | 9.49 | 10.10 |  |
| - Median | 9.10 | 9.53 | 8.87 |  |
| - Range | [3.39, 21.80] | [3.39, 16.93] | [3.49, 21.8] |  |

Any variable was found as statistically different between methylated and unmethylated MGMT groups (p>0.05), suggesting that any of these variables affect to the results of the rest of survival and stratification analyses.

### 3.2. Lack of benefit of temozolomide for MGMT methylated patients with high vascular tumors

#### 3.2.1. Uniparametric Cox regression analysis

Table 2 includes the results of the Uniparametric cox regression analyses for the entire cohort and for the moderate- and high-vascular groups, generated with different proposed cut off thresholds: (I) the threshold proposed in the preliminary study [47], (II) the median HAT rCBVmax of the current study cohort and (III) the threshold calculated with the combination of both cohorts (n = 214 patients).

**Table 2:**
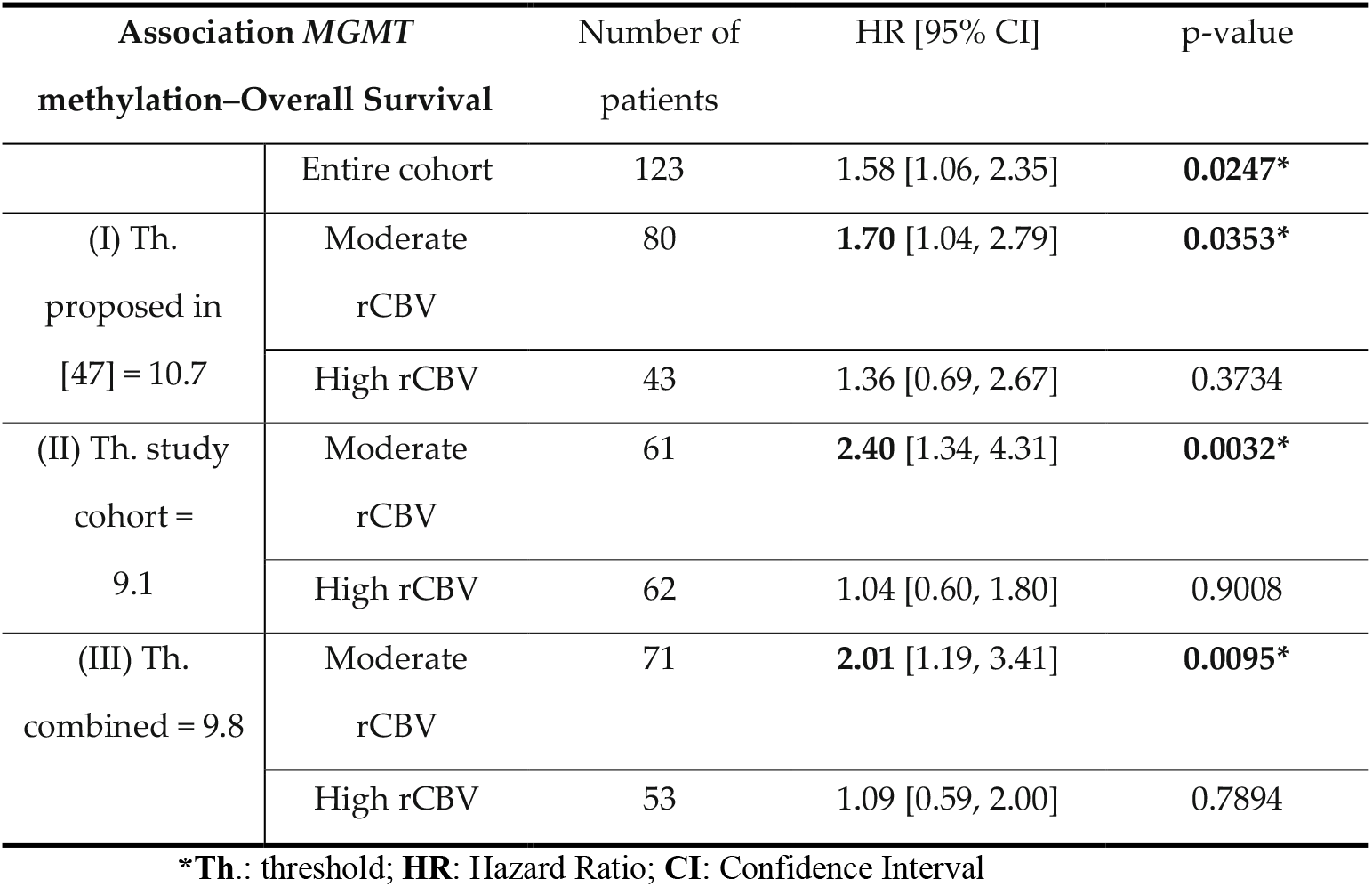
Uniparametric Cox regression results for the entire cohort, and for the moderate- and high-vascular groups, using different proposed cut off thresholds: (I) the threshold proposed in the preliminary study [ref], (II) the median rCBVmax of the present study cohort and (III) the combined threshold calculated with data of both populations (n = 214 patients)

The Uniparametric Cox results show a significant association between the *MGMT* methylation status and patient overall survival (OS) for the entire cohort of 123 patients. However, when this association is analyzed individually for the moderate- and high-vascular groups, we only found significant results for the group of patients with moderate rCBV, regardless of the threshold used. By contrast, we did not find a significant association for the group with high rCBV. These results are repeated for all the vascular groups generated with the three analyzed thresholds, although they are more patent when using the specific threshold of the study cohort, yielding higher HR and lower p-value.

#### 3.2.2. Kaplan Meier and Log rank test

Kaplan Meier results for the entire cohort and for the moderate- and high-vascular groups are included in Table 3.

**Table 3:** Kaplan Meier results for the entire cohort, and for the moderate- and high-vascular groups, generated by the combined cut off threshold (9.8), when comparing the populations with methylated and unmethylated *MGMT*.

|  | Number of patients |  |  | KM results |  |  |  |
| --- | --- | --- | --- | --- | --- | --- | --- |
|  | Total | Meth. *MGMT* | Unmeth. *MGMT* | Median OS Meth. *MGMT* | Median OS Unmeth. *MGMT* | \|ΔOS\| | p-value |
| Entire cohort | 123 | 67 | 56 | 578 | 462 | 114 | **0.0220\*** |
| Moderate rCBV | 71 | 46 | 34 | 641 | 467 | **174** | **0.0129\*** |
| High rCBV | 53 | 21 | 22 | 454 | 461 | 7 | 0.9119 |

\***Meth. MGMT**: methylated *MGMT*; **Unmeth. MGMT**: unmethylated *MGMT*; **OS**: overall survival

The Kaplan Meier results showed significant differences in survival for the entire cohort (p < 0.05) depending on the *MGMT* methylation status. However, these differences in survival time were more significant (lower p-value) and more patent (higher difference in survival days) for the moderate vascular group. For this group we found significant differences (p = 0.0129) in median survival between the populations with methylated *MGMT* and with unmethylated *MGMT* (641 vs. 467 days, respectively), with a difference in OS of 174 days. By contrast, we did not find any difference in survival for the high vascular group, independently of their *MGMT* methylation status.

This differential effect of *MGMT* methylation depending on tumor vascularity is also illustrated in Figure 2, which shows the Kaplan Meier survival curves for each vascular group and for each *MGMT* population. The Kaplan Meier curves using the other two proposed thresholds are included in Figures S1.1 and S1.2 of the Supporting Information.

**Figure 2:**
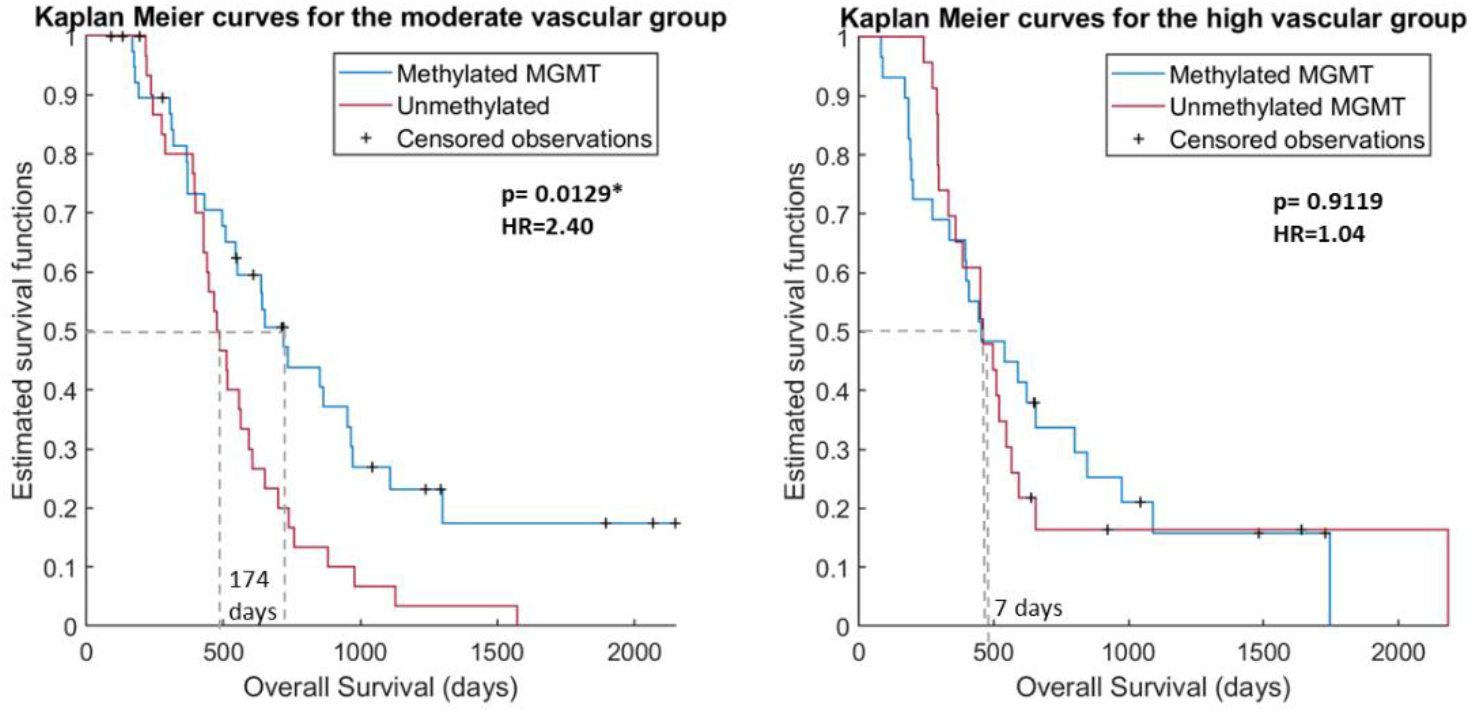
Kaplan Meier curves for the moderate vascular group (left) and for the high vascular group (right) depending on the *MGMT* methylation status.

The Kaplan Meier curves reaffirm the results that the influence of *MGMT* methylation on survival time is only for the moderate vascular group, since only for this group the survival functions are significantly different.

### 3.3. Benefit of adjuvant temozolomide cycles in different groups of glioblastoma patients

#### 3.3.1. Multiparametric Cox regression analysis

Multiparametric Cox results for the entire cohort and, independently for the moderate- and high-vascular groups. including hazard ratios, CIs and p-values are shown in Table 4.

**Table 4:** Multiparametric Cox regression results for the entire cohort, and for the moderate- and high-vascular groups, analyzing the combined correlation between the *MGMT* methylation status and the number of adjuvant Temozolomide cycles with the overall survival.

| Covariables | HR [95% CI] | p-value |
| --- | --- | --- |
| **MGMT** |  |  |
| **Entire cohort** |  |  |
| *MGMT* status | 1.53 [0.96, 2.43] | 0.0727 |
| TMZ cycles | 0.78 [0.70, 0.85] | <0.0001* |
| **Moderate rCBV** |  |  |
| *MGMT* status | 1.75 [1.08, 4.20] | 0.0416* |
| TMZ cycles | 0.78 [0.66, 0.90] | <0.0001* |
| **High rCBV** |  |  |
| *MGMT* | 1.03 [0.56, 1.92] | 0.9121 |
| TMZ cycles | 0.77 [0.68, 0.87] | <0.0001* |

A significant correlation between the number of temozolomide cycles and patient survival was found for the entire cohort, and for the moderate- and high-vascular groups. Nonetheless, only for the moderate vascular group was found a significant correlation for both variables (*MGMT* methylation status and number of temozolomide cycles). These results suggest that the combined effect of these two clinical variables is more relevant for survival time for those patients with moderate tumor vascularity.

Additionally, Figure 3 shows a boxplot per each following group, with survival times depending on the number of adjuvant temozolomide cycles administered:

a. Moderate vascularity + methylated *MGMT*
b. Moderate vascularity + unmethylated *MGMT*
c. High vascularity + methylated *MGMT*
d. High vascularity + unmethylated *MGMT*

**Figure 3:**
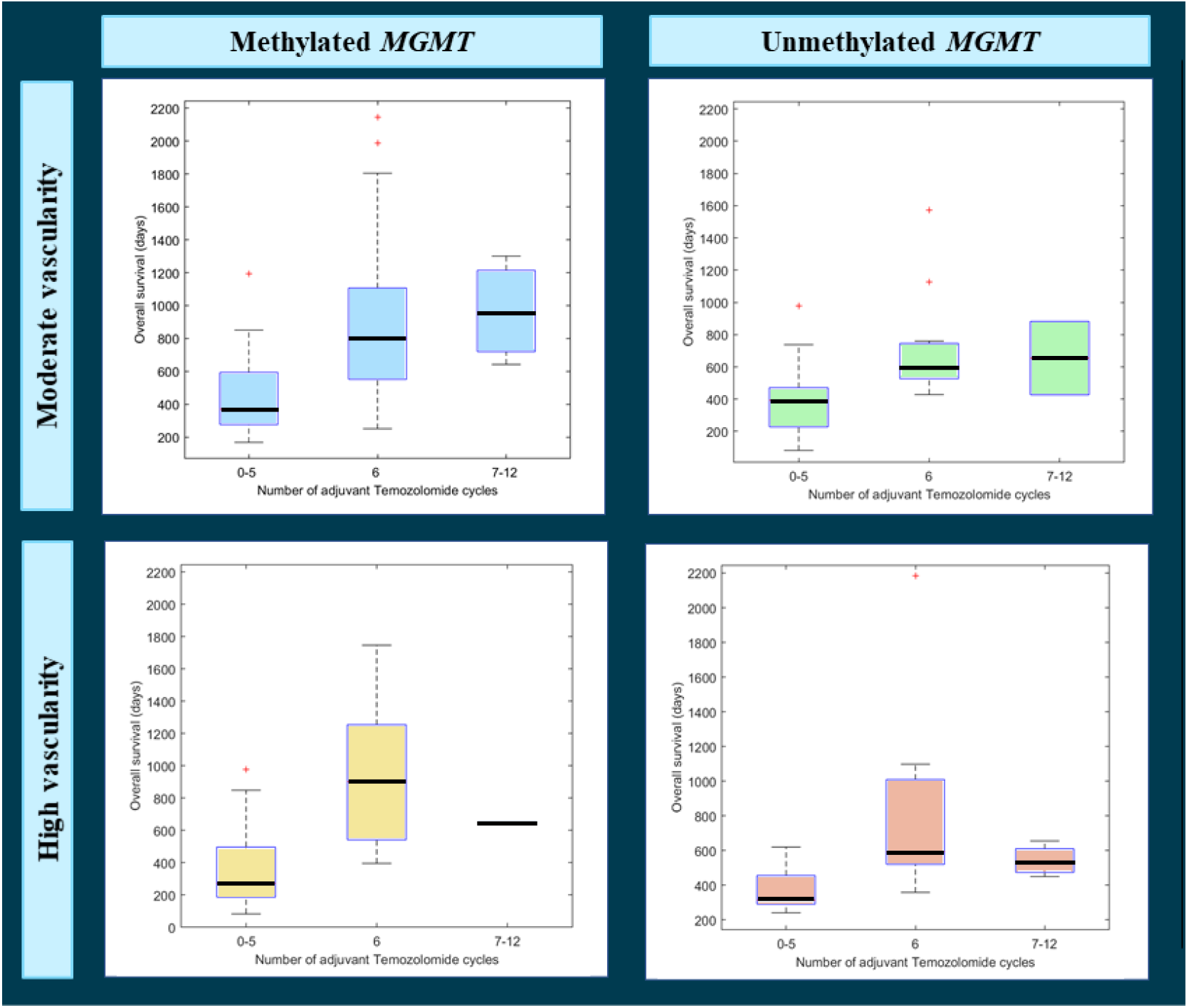
Boxplots analyzing the differences in overall survival according to the administered number of adjuvant Temozolomide cycles (1-5, 6 and 7-12) for different populations: patients with moderate vascularity and methylated *MGMT* (top left), patients with moderate vascularity and unmethylated *MGMT* (top right), patients with high vascularity and methylated *MGMT* (bottom left) and patients with high vascularity and unmethylated *MGMT*.

We can see that for the unmethylated *MGMT* populations (in green and in red), median survival rates do not overcome 700 days in any case, independently from adjuvant temozolomide cycles.

By contrast, different tendencies could be appreciated for the methylated MGMT populations. In the case of patients with moderate vascularity (in blue), median survival rates seem to increase with higher number of temozolomide cycles, being the highest median OS for the group with more than 6 temozolomide cycles.

However, the tendency seems different for the high vascular group (in yellow). Although patients which completed the standard 6-cycle treatment, presented a higher survival rate, to administer more than six cycles do not seem to provide a beneficial effect, even an adverse one.

## 4. Discussion

With the present study we aimed to evaluate the lack of benefit of temozolomide for *MGMT* methylated patients with high vascular glioblastomas, since previous results published in [47] concluded that the combined effect of *MGMT* methylation and moderate vascularity of the tumor causes a benefit in glioblastoma patient overall survival. In addition, the previously proposed threshold has been validated and we propose an upload to be more generalizable in future studies, since it has been calculated with data from 214 patients. Finally, we aimed to investigate the potential benefit of increasing the number of adjuvant temozolomide cycles in different groups of glioblastoma patients according to their MGMT methylation status and tumor vascularity.

To achieve our main purposes, we used an independent and major multicenter cohort of 123 glioblastoma patients. Our results validate the hypothesis proposed in [48], since we have found significant associations (p<0.05) between *MGMT* methylation status and patient survival only for the moderate vascular group of patients, but not for the high vascular group (p>0.05). This is, prognosis of patients with a moderate vascular tumor will be affected by *MGMT* methylation status, while survival times for the high vascular group do not differ, independently of the *MGMT* methylation status. This evidence is also shown when analyzing the Kaplan Meier results: for the moderate vascular group there is a significant difference (p<0.005) of 174 days in median survival depending on presenting methylated or unmethylated *MGMT*, while for the high vascular group there are not significant differences in survival. That is, there is a lack of benefit of temozolomide for *MGMT* methylated patients with high vascular glioblastomas.

Some clinical studies have been developed with the purpose of analyzing the effect of increasing the number of adjuvant temozolomide cycles [14-20], being considered six cycles as the standard [3]. One metanalysis [31] and a retrospective large cohort analysis [32], found no benefits on OS but a possible improvement in Progression free survival. The only randomized phase II trial did not show any benefit in those parameters for the fact of continuing temozolomide for further than 6 cycles. Anyway, this was only a phase II trial with a small number of patients, and it may can be that a particular subgroup of patients get benefit from continuing temozolomide treatment, as our preliminary results suggest.

Considering previous results, which opened the possibility to investigate the different effect of temozolomide in particular subgroups, we explored the benefit of increasing the number of temozolomide cycles depending on their specific *MGMT* status and vascular profile. We investigated the correlation between the number of temozolomide cycles and *MGMT* status for the high- and moderate-vascular groups and we found that only for the moderate vascular group, both variables were significantly associated with patient survival.

Furthermore, we analyzed, in an observational way, the survival patterns of each group (defined by *MGMT* status and vascularity) and with different number of administered temozolomide cycles (<6, 6 or >6). We found specific survival tendencies for each group of patients when administering different number of temozolomide cycles. The group of patients with methylated *MGMT* and moderate vascularity was observed as the only one that benefits from more than 6 temozolomide cycles.

These are preliminary results but considering the interest in deciding more individual treatments for glioblastoma patients, future prospective studies could be relevant to analyze the beneficial effect of providing more than 6 cycles of temozolomide for selected groups of patients. Knowing the marked interpatient heterogeneity, a more personalized approach to treat glioblastoma patients appears to be a potential solution to overcome the heterogeneity and prolonged overall survivals.

The main limitation of this study is the lack of a randomized strategy to provide more than six cycles of adjuvant temozolomide to patients. This is due to the observational and retrospective nature of the study. Assuming that association does not imply causation, our results of analyzing the prognostic effect of temozolomide cycles should be interpreted with caution. However, differences in survival tendencies among groups seem to exist, and future prospective studies could validate these results. These limitations are only referred to the second objective of the study.

## 5. Conclusions

In conclusion, we have demonstrated with a multicentric cohort of 123 glioblastoma patients the lack of benefit of temozolomide for MGMT methylated patients with high vascular tumors. In addition, we have validated the previously proposed threshold (th = 10.7) as useful to stratify patients in terms of vascularity and with significant differences in survival, and we proposed an upload threshold, calculated with both cohorts, (th = 9.8) to be more generalizable in future studies. Finally, we found preliminary results related with the potential benefit of increasing the number of adjuvant temozolomide cycles only for a particular group of patients with MGMT methylation and moderate vascularity, which represents almost a 40% of the study entire cohort. Authors consider clinically relevant a future prospective study analyzing the beneficial effect of providing more than 6 temozolomide cycles in the group of patients with moderate vascularity and methylated MGMT. Positive results could lead us to a more personalized decision making in glioblastoma treatment, allowing prolonged patient survival times.

## Data Availability

No data availability

